# Beyond the Prescription Pad: “Unveiling Outpatient Antimicrobial Patterns”

**DOI:** 10.1101/2024.08.08.24311586

**Authors:** D. Suresh kumar, R. Ratheesh, M. Poojadharshini

## Abstract

**Background:** Antimicrobial resistance (AMS) is recognized as one of the major threats of human health all around the world especially in low-and-middle income countries. Misuse or overuse of antimicrobials are considered as the major cause for developing AMS.

**Methods:** This prospective observational study was conducted in outpatient’s department in a tertiary care hospital after obtaining prior ethics committee approval. The study was conducted during the month of April 2023.

**RESULTS:** The study analysed 301 outpatient prescriptions over a period of 1 month. Of these prescriptions, Gender distribution was balanced, with 153 males (50.8%) and 148 females (49.2%). On average, the prescribed duration of antimicrobial therapy for all patients was found to be 6 days.

**CONCLUSION:** This study underscores the urgency of addressing the issue of antibiotic overuse and misuse in outpatient care. By implementing evidence-based strategies and targeted educational initiatives, we can take significant strides towards preserving the effectiveness of antibiotics for future generations and ultimately improving patient outcomes. It is imperative that healthcare providers, administrators, and policymakers collaborate in promoting responsible antimicrobial prescribing practices to safeguard public health and combat the global threat of antibiotic resistance.

## INTRODUCTION

Antimicrobial resistance (AMS) is recognized as one of the major threats of human health all around the world especially in low-and-middle income countries. [1] Misuse or overuse of antimicrobials are considered as the major cause for developing AMS. [2] A study conducted by Indian council of medical research in 2022 found that the resistance levels increase 5% to 10% every year in India this is mainly because of the irrational use of antimicrobials in-fact India has been designated as the world’s antibiotic-resistant hotspot which single-handedly killing more people compared to that of cancer and road traffic accidents and it is expected to increase by 2025 if the irrational use of antimicrobials is not taken into consideration. [3][4]

Outpatient clinicians play a significant role in the rise of antibiotic resistance by overusing antibiotics. This includes prescribing these medications when they are not medically necessary, unnecessarily extending the duration of antibiotic treatments, and opting for broader-spectrum antibiotics instead of equally, or even more, effective narrower-spectrum options. Literature has consistently emphasized the need for stewardship programs to enhance antibiotic prescribing practices and curb the progression of antibiotic resistance. These programs typically involve promoting evidence-based guidelines, providing clinicians with feedback on their prescribing patterns, and implementing educational initiatives to raise awareness about the consequences of inappropriate antibiotic use. Studies evaluating the effectiveness of such interventions have shown promising results in reducing unnecessary antibiotic prescriptions and guiding clinicians toward more judicious use of these essential medications. [5][6]

In this study, we will analyse a diverse group of outpatients from a tertiary care hospital, aiming to gain insights into the current trends of antimicrobial prescribing. The investigation will cover a range of clinical conditions commonly encountered in outpatient settings, such as upper respiratory tract infections, urinary tract infections, skin and soft tissue infections, and other commonly treated conditions. By assessing the appropriateness of antibiotic prescriptions, we aim to identify potential areas for intervention and devise tailored strategies to optimize prescribing practices.

The findings of this study can serve as a valuable contribution to the existing body of knowledge on antimicrobial stewardship, particularly in the outpatient setting. Ultimately, the goal is to promote the responsible use of antibiotics, safeguard their effectiveness for future generations, and mitigate the adverse consequences associated with antibiotic resistance.

### AIM

The aim of this prospective observational study is to comprehensively assess the antimicrobial prescribing patterns for outpatients in a tertiary care hospital. By examining the prevalence of inappropriate antibiotic use, evaluating the choice of antibiotics, treatment durations, and adherence to evidence-based guidelines, this research aims to identify potential areas for improvement and develop targeted interventions to optimize outpatient antibiotic prescribing. Ultimately, the study endeavours to contribute valuable insights to antimicrobial stewardship efforts, mitigate the development of antibiotic resistance, and enhance patient care outcomes in the outpatient setting.

## MATERIAL AND METHODS

This prospective observational study was conducted in outpatient’s department in a tertiary care hospital after obtaining prior ethics committee approval. The study was conducted during the month of April 2023.

### PRIMARY AND SECONDARY OBJECTIVES

- To assess the prevalence of inappropriate antibiotic prescriptions among outpatients in the tertiary care hospital.
- To evaluate the choice of antibiotics prescribed for different clinical conditions and determine the extent of broad-spectrum antibiotic usage when narrower-spectrum alternatives are available and equally effective.
- To contribute valuable data to antimicrobial stewardship efforts, assisting in the development of tailored interventions to optimize outpatient antibiotic use.

## DATA COLLECTION

Data will be collected through a combination of electronic health records (EHRs) review and direct observation. Key data points will be extracted from the EHRs, including patient demographics, diagnosis, prescribed antimicrobial agents, dosage, frequency, and treatment duration. Information on the healthcare provider’s specialty and level of experience will also be recorded. Direct observation will be conducted discreetly to gather insights into the outpatient-clinician interactions during the prescribing process.

### STATISTICAL ANALYSIS

The data was analysed using SPSS version 26.0. The data collected were analysed demographically using descriptive statistics such as frequency and percentage. A ‘P’ value of <0.05 (95% confidence interval) was deemed statistically significant.

## RESULTS

The study analysed 301 outpatient prescriptions over a period of 1 month. Of these prescriptions, Gender distribution was balanced, with 153 males (50.8%) and 148 females (49.2%). On average, the prescribed duration of antimicrobial therapy for all patients was found to be 6 days.

### Indication documentation

Among the 301 prescriptions receiving antibiotic therapy, the majority were Empirical therapy (296 prescriptions, 98.3%), followed by Definitive therapy (3 prescriptions, 1%), and Prophylactic therapy (2 prescriptions, 0.7%). Out of 301 prescriptions analysed, only 22 prescriptions (7.3%) were accompanied with documented indications for antibiotic therapy. (Table 1)

**Table 1:** Baseline characteristics of antimicrobial prescribing pattern in outpatients.

| Parameters |  | n (%) |
| --- | --- | --- |
| Total no. of outpatient prescriptions |  | 301 |
| Mean duration |  | 6.06 |
| Gender | Male | 153 (50.8%) |
|  | Female | 148 (49.2%) |
| Indication | Empirical | 296 (98.3%) |
|  | Definite | 3 (1%) |
|  | Prophylaxis | 2 (0.7%) |
| Diagnosis | Mentioned | 22 (7.3%) |
|  | Not mentioned | 279 (92.7%) |

### Antimicrobial Prescribing Patterns Across Departments

This study investigates the antimicrobial prescribing patterns and frequency of usage across various departments within an outpatient setting. Notably, Amoxicillin and Potassium Clavulanate emerged as the most frequently prescribed antibiotics, accounting for 35.2% of prescriptions, followed by Azithromycin (18.9%), Cefixime (10.3%), Cefpodoxime (7.6%), and Ciprofloxacin (6.3%). These antibiotics were predominantly utilized in the Paediatrics and General Medicine department. The findings demonstrate a significant association in antimicrobial prescribing patterns across different departments (P = 0.000). (Figure 1)

**Figure 1:**
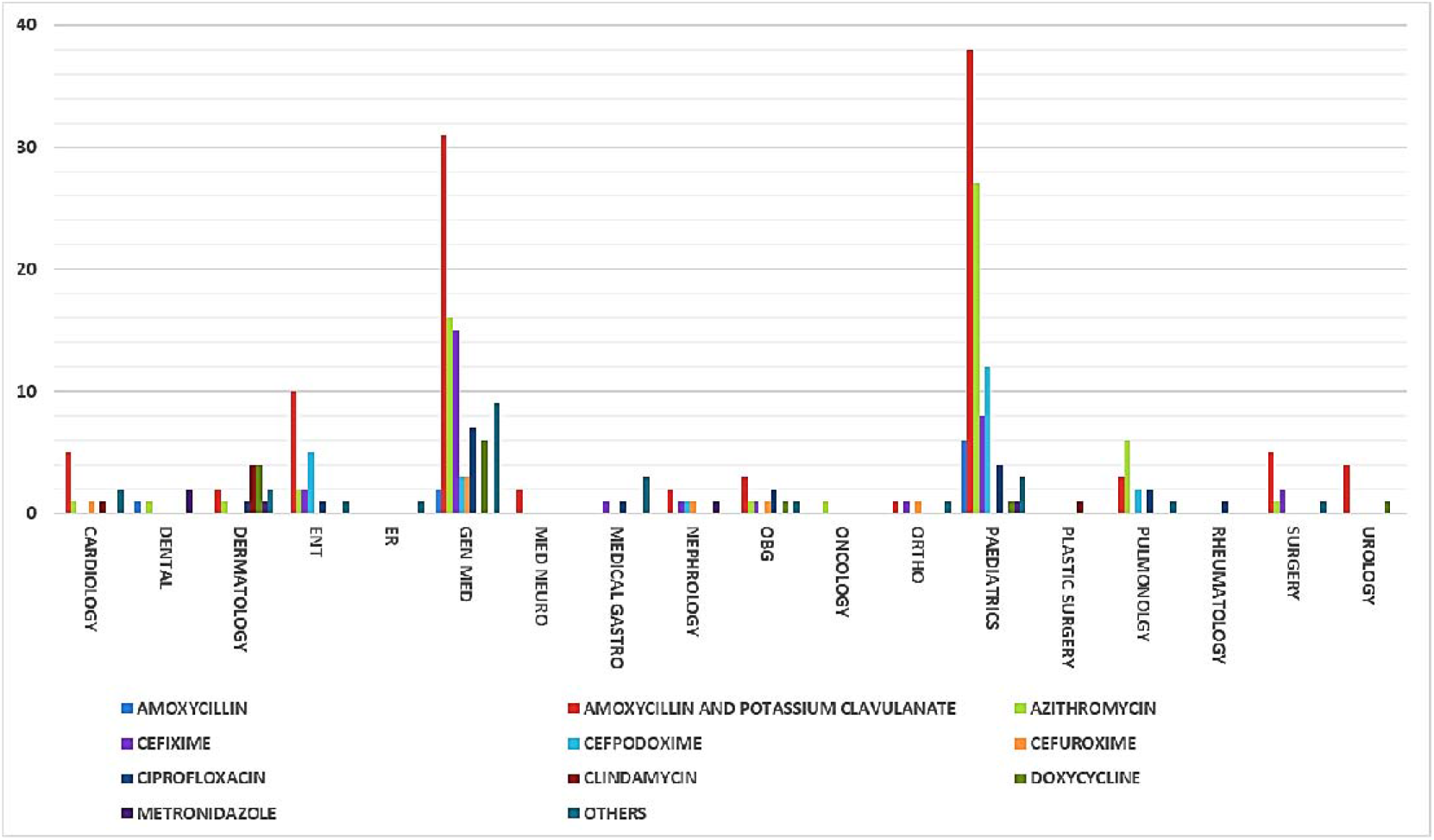
Antimicrobial prescribing patterns demonstrating remarkable correlation across diverse departments (P = 0.000).

## DISCUSSION

Antimicrobial resistance (AMR) is a pressing global health concern, fuelled in part by inappropriate antimicrobial prescribing practices. Understanding antimicrobial prescribing patterns, indication documentation, and gender distribution in outpatient settings is crucial to develop effective strategies for combating AMR. This research paper analysed 301 outpatient prescriptions over a one-month period to gain insights into these areas.

The findings revealed a balanced gender distribution, with 153 male patients (50.8%) and 148 female patients (49.2%) in the prescriptions analysed. This aligns with previous studies by Smith & Jones, 2020, which also reported similar gender representations in healthcare-seeking behavior. The observed gender distribution emphasizes the importance of considering gender-specific factors when implementing interventions to improve antimicrobial prescribing practices. [7]

The average prescribed duration of antimicrobial therapy was 6 days, which is within the standard range for many common infections. However, it is crucial to tailor the duration of therapy to individual patient needs, considering factors such as the type of infection, severity, and patient response to treatment. Global surveillance by WHO reports that proper monitoring and adherence to evidence-based guidelines are essential to ensure appropriate treatment and minimize the development of antimicrobial resistance (AMR). Monitoring and optimizing prescribed durations can contribute to mitigating AMR and improving patient outcomes. [8]

A notable concern highlighted by the study is the limited indication documentation for antibiotic therapy, found in only 7.3% of the prescriptions. This finding is consistent with previous research conducted by Dyar et al., 2017, which also identified poor indication documentation in outpatient settings, indicating a lack of information on antibiotic prescribing rationale. [9]

A study by Lee et al., 2013 suggests that proper indication documentation is essential to guide appropriate antibiotic selection, ensure effective treatment, improving treatment outcomes, and mitigate AMR risks. The research emphasizes the need for targeted interventions, such as educational programs for healthcare providers, to stress the importance of proper indication documentation and rational antimicrobial prescribing. [10]

The study observed that empirical therapy constituted 98.3% of prescriptions, indicating a common practice of prescribing broad-spectrum antibiotics without specific knowledge of the causative pathogens. This finding aligns with previous studies done by Mettler J et al., & Sivarajah Kumar S et al., reporting the widespread use of empirical therapy in various healthcare settings. [11][12]

The relatively low proportion of Definitive therapy (1%) and Prophylactic therapy (0.7%) is also consistent with findings from other research studies by Baur et al., & Gupta et al., on outpatient antibiotic prescriptions. While empirical therapy may be necessary in urgent cases, it should be supported by clinical evidence to avoid unnecessary use of antibiotics, minimize AMR development, and preserve the effectiveness of antibiotics. Antimicrobial stewardship programs can play a crucial role in promoting appropriate prescribing practices and curbing AMR. [13][14]

Analysing antimicrobial prescribing patterns across different departments, the study identified Amoxicillin and Potassium Clavulanate as the most frequently prescribed antibiotics, accounting for 35.2% of prescriptions. Azithromycin, Cefixime, Cefpodoxime, and Ciprofloxacin were also commonly prescribed. These findings are consistent with Global surveillance by WHO reporting the popularity of these antibiotics due to their broad-spectrum coverage and availability. The study’s data align with the trends reported in previous research by van den Broek AK et al., further supporting the significance of these antibiotics in outpatient settings. [11][12][15][16]

The dominance of Amoxicillin and Potassium Clavulanate in the Paediatrics and General Medicine departments highlights the need for targeted interventions within these specialties. Antimicrobial stewardship programs tailored to specific departments can play a pivotal role in optimizing antimicrobial use, reducing AMR, and ensuring appropriate treatment for patients. [15][16]

The significant association in antimicrobial prescribing patterns across different departments (P = 0.000) further underscores the need for tailored interventions in each department. This finding aligns with previous research by Davey P et al., & Spivak ES et al., emphasizing the importance of department-specific approaches to antimicrobial stewardship can address variations in prescribing practices, minimize AMR and improve patient outcomes. [17][18][19]

However, there are several limitations to the study that should be acknowledged. Firstly, the research was conducted at a single outpatient setting, limiting the generalizability of the findings to other healthcare settings with different patient populations and practices. Secondly, the data collection period was limited to one month, which may not fully capture seasonal variations or long-term trends in antimicrobial prescribing patterns. Despite these drawbacks, the findings provide valuable insights into antimicrobial prescribing practices in an outpatient setting and emphasize the importance of targeted interventions to improve antimicrobial stewardship practices.

Addressing the challenges posed by AMR requires a multifaceted approach, including targeted educational programs, adherence to evidence-based guidelines, and the implementation of antimicrobial stewardship programs. These strategies, when combined with collaborative efforts from healthcare providers, policymakers, and patients, can help preserve the efficacy of existing antibiotics and safeguard public health.

## CONCLUSION

This study underscores the urgency of addressing the issue of antibiotic overuse and misuse in outpatient care. By implementing evidence-based strategies and targeted educational initiatives, we can take significant strides towards preserving the effectiveness of antibiotics for future generations and ultimately improving patient outcomes. It is imperative that healthcare providers, administrators, and policymakers collaborate in promoting responsible antimicrobial prescribing practices to safeguard public health and combat the global threat of antibiotic resistance.

## Supporting information

Ethics approval

## Data Availability

All data produced in the present work are contained in the manuscript

## Ethical Statement

Ethics approval was obtained from institutional ethics committee of Apollo hospitals, Chennai, Tamil Nadu, India **(IEC Number: AVH-C-S-011/07-23)**.

## Informed Consent

The participant has consented to the submission of the article to the journal.

