## Supplementary material for "Beyond the Prescription Pad: “Unveiling Outpatient Antimicrobial Patterns”": Ethics approval

NEW EC Reg. No. - EC/NEW/INST/2022/TN/0195

25 July 2023

**To,**
Dr. Suresh Kumar,
Principal Investigator,
Apollo Speciality Hospital, Vanagaram,
Chennai, Tamil Nadu, India.

**Ref: IEC-BMR App No.:** AVH-C-S-011/07-23 **Protocol No.:** NA

**Title:** A Prospective observational study on assessing the Antimicrobial Prescribing patterns for outpatients in a tertiary care hospital.

**Sub: Final Approval** (Subsequent to your Ltr. Dt.27 June 2023).

Dear Dr. Suresh Kumar,

The Institutional Ethics Committee- Bio Medical Research, Apollo Hospitals, Chennai reviewed and discussed the documents submitted by you related to the conduct of the above referenced study at its meeting held on 18th July 2023.

**Documents submitted:**

| Sl. No. | Contents | Version and Date |
|---|---|---|
| 1 | Study Proposal | NA |
| 2 | CV & MRC of PI | NA and Dated 27 Jun 2023 |
| 3 | Questionnaire Form | NA |
| 4 | Wavier of Consent | NA and Dated 27 Jun 2023 |

The following Institutional Ethics Committee – Bio Medical Research members were present at the meeting held on 18th July 2023 at 02.30 pm.

| S. No. | Name | M/F | Qualification | Affiliated to Institution (Y/N) | Designation | Position In The Committee |
|---|---|---|---|---|---|---|
| 1 | Dr. R. Ramakrishnan | M | M.Sc., Ph.D, M.A.E | No | Consultant/ Ex -Scientist "G | Chairperson |
| 2 | Dr. Hemalatha | F | MBBS | Yes | Medical Superintendent | Member Secretary |
| 3 | Dr. M. Nirumal Rakkesh | M | MBBS, M.D. (Pharmacology) | Yes | Pharmacologist | Basic Medical Scientist |

| | | | | | | |
|---|---|---|---|---|---|---|
| 4 | Dr. K. Sathyamurthi | M | M.A. (Social Work), PGDHRM, Ph.D | No | HOD Department of Research Programme in Social Work | Member (Social Scientist) |
| 5 | Ms. Swapna Sundar | F | BA, BL, ML, M. Phil Law | No | CEO, IP Dome IP Strategy Advisors Pvt. Ltd. | Member (Lawyer) |
| 6 | Dr. Kirubakaran | M | MBBS, MD | Yes | Consultant Cardiologist | Clinician |
| 7 | Mrs. Padma Priya | F | B.A. (History) & M.A. (Hindi) | No | Home Maker | Layperson |

- *No Member had any conflict of interest and participated in the decision making process.*

After due ethical and scientific consideration, the Ethics Committee has approved all the documents and the study to be conducted by you in the presented form.

The Ethics Committee should also be informed about the progress of the study on **Half Yearly basis.** Any changes in the protocol and patient information/ informed consent should be submitted for review and approval. Submit a report of protocol deviations/ violations and serious adverse event as per regulatory timeline and mention the reason for delay, if any.
A copy of final clinical study report should be submitted.

**Please note the period of validity of this approval is for one calendar year and ends on 24 July 2024.**

The Institutional Ethics Committee – Bio Medical Research is constituted and works as per ICH-GCP, National Ethical Guidelines for Biomedical and Health Research involving Human Participants (ICMR 2017) and New drugs and Clinical Trial Rules, March 2019.

Yours Sincerely,

MEMBER SECRETARY
INSTITUTIONAL ETHICS COMMITTEE BIO MEDICAL RESEARCH
APOLLO HOSPITALS, AHEL
CHENNAI, TAMILNADU.

**Dr. Hemalatha**
Member Secretary
Institutional Ethics Committee – Bio Medical Research
Apollo Hospitals, Chennai
Tamil Nadu, India.

**IEC-BMR Application No.:** AVH-C-S-011/07-23 (NOTE: Please quote this application no. in all your future communications).

**Status: Approved**
